# Late-Ensemble of Convolutional Neural Networks with Test Time Augmentation for Chest XR COVID-19 Detection

**DOI:** 10.1101/2022.02.25.22271520

**Authors:** Abdul Qayyum, Imran Razzak, Moona Mazher, Domenec Puig

**Affiliations:** University of Burgundy, Dijon, France; Deakin University Geelong, Australia; University Rovira i Virgili, Spain

**Keywords:** COVID-19, Chest XR COVID-19, COVID-19 Challenge, Ensemble, X-Ray

## Abstract

COVID-19, a severe acute respiratory syndrome aggressively spread among global populations in just a few months. Since then, it has had four dominant variants (Alpha, Beta, Gamma and Delta) that are far more contagious than original. Accurate and timely diagnosis of COVID-19 is critical for analysis of damage to lungs, treatment, as well as quarantine management [7]. CT, MRI or X-rays image analysis using deep learning provide an efficient and accurate diagnosis of COVID-19 that could help to counter its outbreak. With the aim to provide efficient multi-class COVID-19 detection, recently, COVID-19 Detection challenge using X-ray is organized [12]. In this paper, the late-fusion of features is extracted from pre-trained various convolutional neural networks and fine-tuned these models using the challenge dataset. The DensNet201 with Adam optimizer and EffecientNet-B3 are fine-tuned on the challenge dataset and ensembles the features to get the final prediction. Besides, we also considered the test time augmentation technique after the late-ensembling approach to further improve the performance of our proposed solution. Evaluation on Chest XR COVID-19 showed that our model achieved overall accuracy is 95.67%. We made the code is publicly available^1^. The proposed approach was ranked 6th in Chest XR COVID-19 detection Challenge [1].

## I. Introduction

COVID-19 is an infectious disease caused by the SARS-CoV-2 virus that shares similarities with SARS and MERS viruses that were previously reported in 2003 and 2012. It is ongoing life threatening virus that the world has been facing since 2019. As of October 2021, COVID-19 has four dominant variants spreading among global populations: Alpha /B.1.1.7, first found in UK (London and Kent), Beta /B.1.351 found in South Africa, Gamma/P.1 found in Brazil Variant and the Delta/B.1.617.2 found in India Variant. Among these, Delta is one of the most contagious and deadly that has comparatively high high infectivity and extreme lethality. Up to 30 October 2021, 246 million COVID cases have been diagnosed, with 4.99M deaths worldwide. United States and India are the two leading countries that affected most with 45.9M/0.745M and 34.2/0.457M with cases/death. Figure 1^2^ shows the COVID-19 cases in France and Australia on 7th October 2021. Recently, deep learning has been widely applied in health especially in analyzing the images i.e. Chest images (MR, CIT and Ultrasound) for COVID image analysis [3], [4], [8], [13]. Qayuim et al. presented depth-wise neural network for COVID-19 diagnosis using CT image [10], [11]. They have uniformly scale all the dimensions and performed multilevel feature embedding result in increase feature representation. ConvNets upscaling is considered by balancing the scaling factor. Depth wise multilevel concatenation is applied to combine different features representation levels that improve diagnostic performance. In another work, Wang et al. proposed noise robust dice loss approach for lesion segmentation ensemble networks (COPLE-Net) for the segmentation of COVID19 [13]. Muhammad et al. presented a convolutionary neural network with much lower number of learning parameters. The network has five main layers of convolution connectors with multi-layer fusion functionality in each block, resulting in detection performance. Experiments on a publicly dataset showed 92.5% precision and 91.8% accuracy [6]. Wang et al. applied self-created CNN to learn individual image-level representations, and rank-based average pooling and multiple-way data augmentation are applied to improve the diagnostic performance [14].

**Fig. 1.**
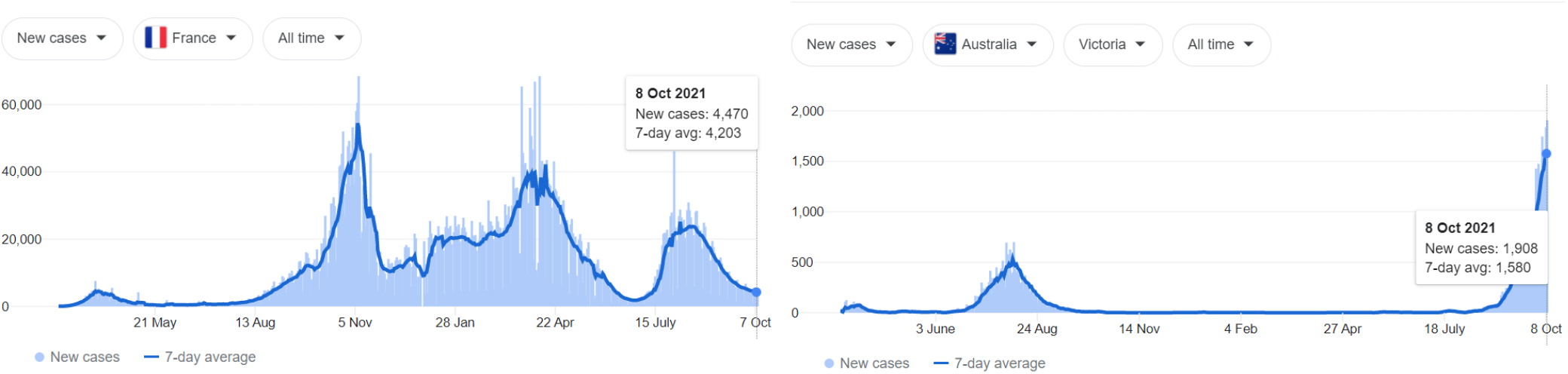
Growth of COVID-19 Cases in France and Australia: Spikes in COVID cases shows COVID-19 1st, 2nd and 3rd wave

GNN Graph neural network is deployed to learn relation-aware representations, and deep feature level fusion is performed to fuse individual image-level features and relation-aware features from both CNN and GNN. Hussain et al. applied CNN named CoroDet for COVID-19 diagnosis using raw chest X-ray and CT. CoroDet showed efficient diagnostic performance in differentiating COVID, and Normal pneumonia patient [5]. CorDet showed a significant gain in performance and achieved 99.1%, 94.2% and 91.2$ classification accuracy for for 2 class, 3, and 4 class classification. Abbas et al. presented DeTraC (Decompose, Transfer, and Compose) for the classification of COVID patient [2] as a results, the model is able to deal with any irregularities by first investigating the boundary of the target class. Results showed that DeTraC achieved 93.1% and 100%sensitivity for COVID-19 diagnosis. In another work, Zebin et al. used multiple pre-trained models as feature extractors and achieved of 90%, 94.3%, and 96.8% over all accuracy using the VGG16, ResNet50, and EfficientNetB0, respectively [16]. Besides, the generative adversarial framework is also utilized to deal with minority COVID-19 class. Phasm fine-tuned alexNet, GoogleNet, and SqueezeNet without data augmentation for 2-class and 3-class COVID-19 diagnosis [9]. Wang et al. presented a computer aided diagnostic framework that consists of Discrimination-deep learning and Localization-deep learning [15]. Discrimination-deep learning extracts lung features from chest X-ray images followed by training. Followed by Localization-deep learning which was trained with 406-pixel patches to extract the recognized of X-ray images and classify them to left lung, right lung or bipulmonary. X-ray. The two-layered approach showed better performance than radiologists and achieved 98.71% and 93.03 % accuracy, classification and localization, respectively. To provide effective and efficient COVID-19 diagnosis, one of the vital effort is Chest XR COVID-19 Chest X-ray image classification Challenge. In this paper, we presented late ensemble convolutional neural networks with test time augmentation for detecting COVID-19 affected patient using chest X-ray images. Experiments were conduction on the Chest XR COVID-19 detection challenge dataset. We have used proposed models trained weights to average the features in post ensemble using mean average voting technique and then passed to the Test Time augmentation technique. Evaluation of the test dataset showed that our model had achieved significantly better performance.

## II. Methodology: Ensemble Network

In this section, we present the proposed deep learning framework for COVID-19 Diagnosis. Figure 2 illustrate the proposed ensemble framework. To improve the diagnostic performance, we have trained various pretrained models such as DensNet121, DensNet161, DensNet201, ResNet18,ResNet34, EfficienNet-b0-B7, ResNext50, ResNext101 and MobileNetV2. We have fine-tuned the weights of each model using the Challenge dataset based on a pre-trained convolutional neural network. We have selected best-performing methods and performed late ensembling. To improve the performance, we applied different data augmentation (horizontalFlip and verticalFlip). Finally, we have used models trained weights to average the features in post ensemble using mean average voting technique and then passed to the Test Time augmentation technique. Similar to what data augmentation is doing to the training set, the purpose of test time augmentation is to perform random modifications to the test images. Thus, instead of showing the regular, “clean” images only once to the trained model, we show the augmented images several times and then average the predictions of each corresponding image and take that as the final prediction. We used horizontal and vertical flipping as test time augmentation in our proposed solution and received fruitful results.

**Fig. 2.**
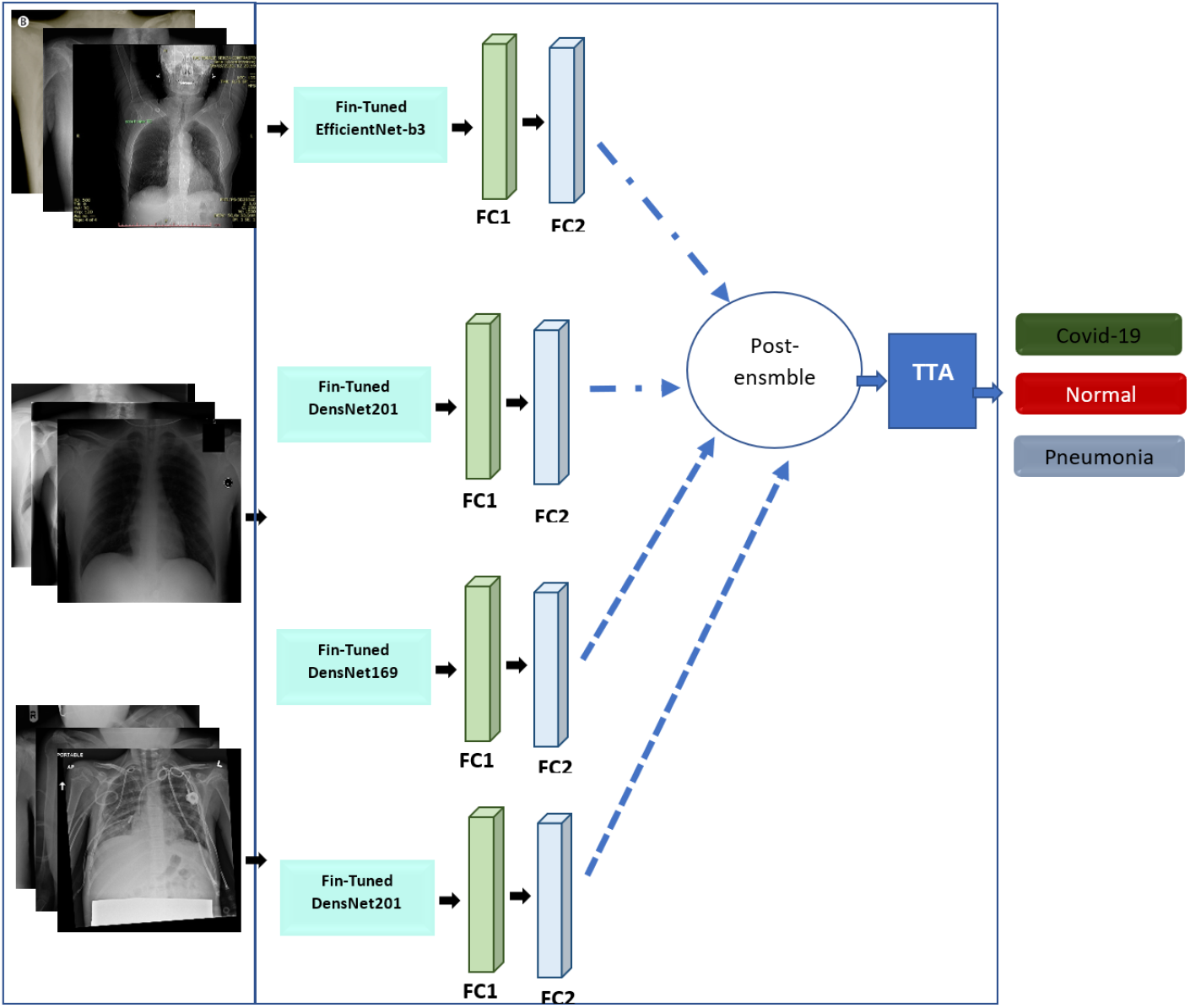
Proposed Late-Ensemble Network

## III. Results and Discussion

In this section, we provide the dataset, parameter and experimental setup, and results. In order to compare the performance, we have used accuracy, sensitivity, and specificity.

### A. Dataset

Int this experiment, we have used Chest XR COVID-19 detection challenge dataset [1]. Chest XR COVID-19 is a large-scale multiclass (COVID-19, Normal and Pneumonia) Chest X-ray image classification dataset. Figure 3 shows COVID-19, Pneumonia and healthy images from dataset. The dataset consist of 20,000+ images which are divided into train (17,955 images) and validation (3,430 images) sets.

**Fig. 3.**
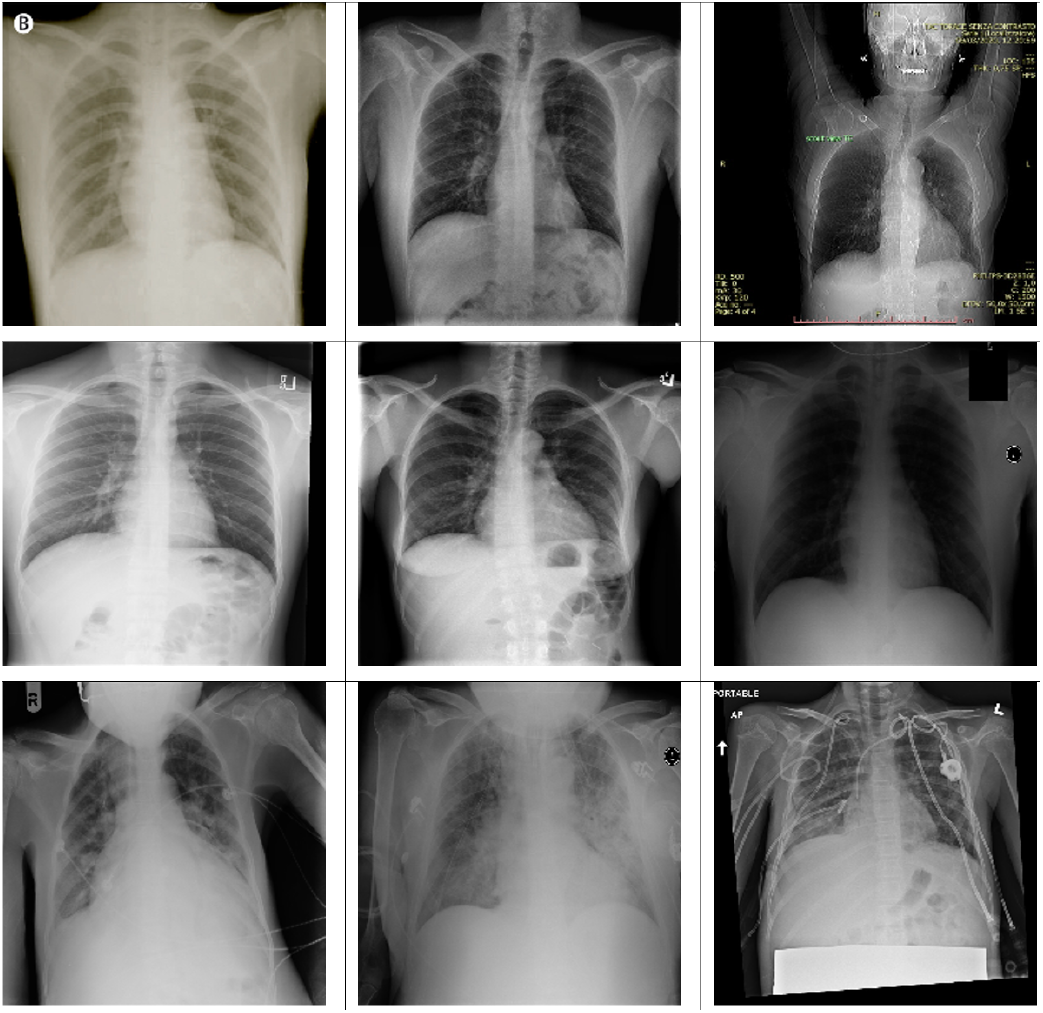
First row-COVID-19 samples, second row-Normal and third rowPneumonia

### B. Parameters

In this experiment, we have used PyTorch library for model development. The proposed model was trained with different hyperparameters. We set the learning rate to 0.0001 with Adam, adamx, adamW optimizers. As the dataset is imbalanced classes samples, the weighted cross-entropy function is used as a loss function between the output of the model and the ground-truth sample. The inverse class frequencies for weight balancing have been used to calculate the weighted crossentropy loss function. The higher-class samples require less weight and fewer class samples need more weight values. The 32 batch-size with 500 epochs has been used with 15 early stopping steps. The best model weights have been saved for prediction in the validation phase. The 224×224 input image size was used for training and prediction. We have used the V100 tesla NVidia-GPU machine for training and testing the proposed model. The detail of the training protocol is described in I.

### C. Discussion

In this paper, we have fine-tune and performed late ensemble of state of the art performing networks such as DensNet121, DensNet161, DensNet201, ResNet18,ResNet34, EfficienNet-b0-B7, ResNext50, ResNext101 and MobileNetV2. Experiments were conducted on the Challenge XR dataset. Table III shows the performance of different methods. Notice that we have ensembled different combinations of networks (independent efficientNet, DenseNet, and ensemble of two, three, and four networks. To improve the performance, we applied different data augmentation (horizontalFlip and verticalFlip). Figure 4 shows the feature extracted from last fully connected layers for three classes. We used TSNE to plot high dimensions features into 2-dimensional. We can observe that ensembles of four networks (Figure (a)) showed that very few features overlap comparatively with other class features.

**TABLE I.** Network Parameters

|  |  |
| --- | --- |
| Data augmentation methods | HorizontalFlip (p=0.5), VerticalFlip (p=0.5), RandomBrightnessContrast (p=0.8) |
| Initialization of the network | “he” normal initialization |
| Batch size | 32 |
| Patch size | 224x224 |
| Total epochs | 500 |
| Optimizer | Adam, Adamx, AdamW |
| Initial learning rate | 0.0001 |
| Learning rate decay schedule | None |
| Stopping criteria, and optimal model selection criteria | The stopping criterion is reaching the maximum number of epoch |

**TABLE II.** Comparative Evaluation of Different Network on Validation Set

| Models | Accuracy | Sensitivity | Specificity |
| --- | --- | --- | --- |
| Ensemble-four models with TTA | 0.9687 | 0.968 | 0.9691 |
| Ensemble-three models with TTA | 0.9662 | 0.9613 | 0.9687 |
| Ensemble-two models with TTA | 0.9644 | 0.9617 | 0.9633 |
| Ensemble-two without TTA | 0.9613 | 0.9610 | 0.9641 |
| EfficientNet-b3 | 0.9562 | 0.9560 | 0.9565 |
| DensNet201(adam optimizer) | 0.9567 | 0.9558 | 0.9542 |
| RensNet201(adamax optimizer) | 0.9513 | 0.9527 | 0.9545 |
| DensNet201 | 0.9412 | 0.9413 | 0.9421 |

**TABLE III.** Comparative Evaluation of Different Network on Test Set

| Models | Accuracy | Sensitivity | Specificity |
| --- | --- | --- | --- |
| Ensemble-four models with TTA | **0.9567** | **0.9567** | **0.9577** |
| Ensemble-three models with TTA | 0.9558 | 0.9558 | 0.9563 |
| Ensemble-two models with TTA | 0.9558 | 0.9558 | 0.9559 |
| Ensemble-two without TTA | 0.9525 | 0.9525 | 0.9539 |
| EfficientNet-b3 | 0.9450 | 0.9450 | 0.9455 |
| DensNet201(adam optimizer) | 0.9458 | 0.9458 | 0.9462 |
| RensNet201(adamax optimizer) | 0.9475 | 0.9475 | 0.9484 |
| DensNet201 | 0.9300 | 0.9300 | 0.9308 |

**Fig. 4.**
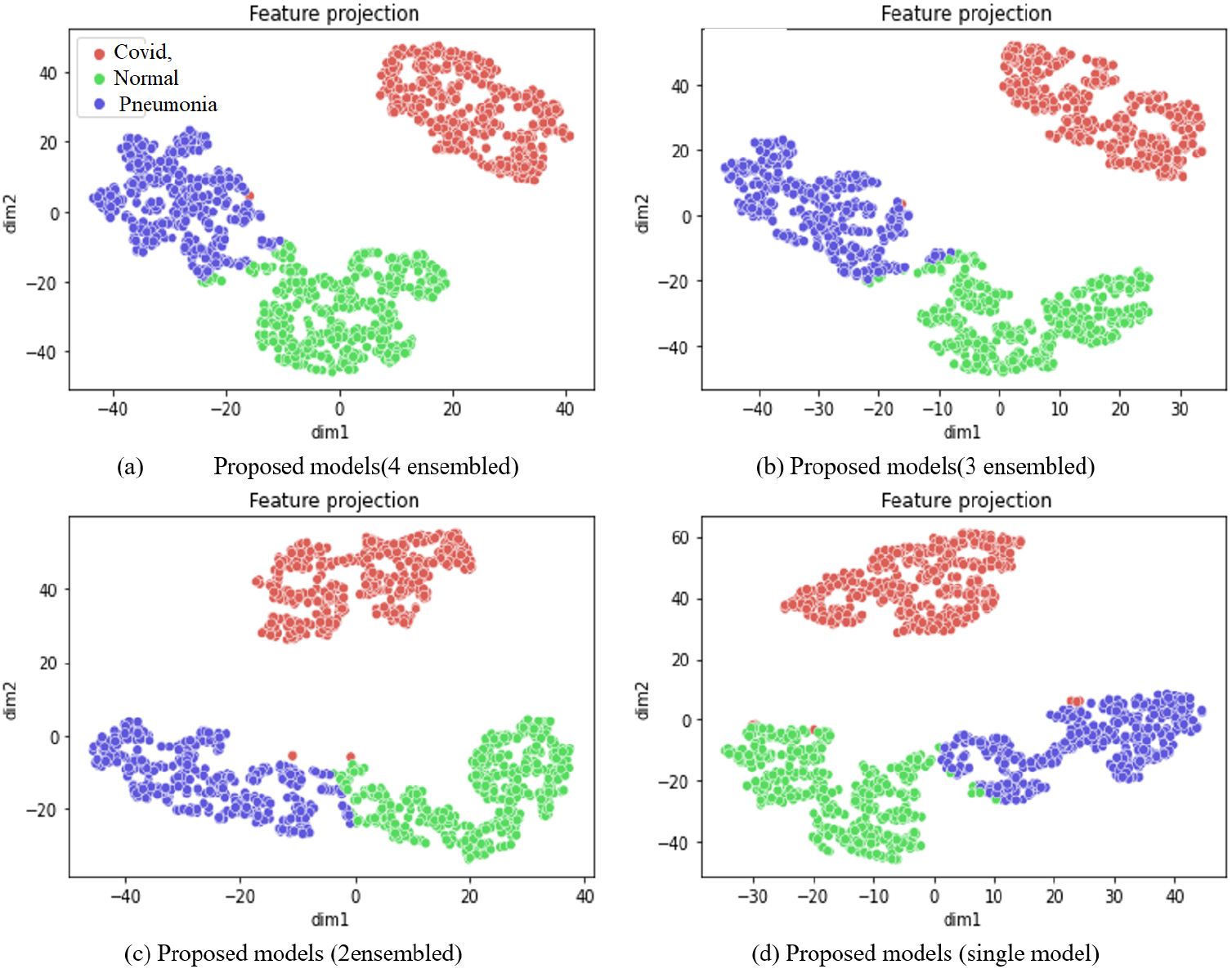
Feature projection of logits layer on test dataset for three classes using proposed models. Class 0 represented covid, 1 represnted normal and 2 represented Pneumonia

Experimental results showed that the ensemble of four networks showed 95.67%, 95.67%, and 95.77% accuracy, sensitivity and specificity, respectively. We can observe that the combination of three models shows slightly lower performance than the ensemble of three network (95.58%, 95.58%, and 95.63% respectively). Similar trend can be noticed with single network. We can observe that EfficientNet and DenseNet showed similar performance. The individual DensNet201 with admax optimzer produced 0.9475 accuracy as compared to single DensNet201 with adam optimizer. To improve the performance, we applied different data test time augmentation, which helped to improve the performance slightly. The proposed solution showed high accuracy on the test dataset; however, it is computational complex in prediction due to test time augmentation. In the future, we will try to reduce the test or inference time by either reducing the parameters of the proposed model.

## IV. Conclusion

The exponential growth in COVID cases is overwhelming health-care system. Every patient with respiratory illness cannot be tested using conventional techniques with limited testing kits. This paper presented late ensemble convolutional neural networks with test time augmentation for detecting COVID-19 affected patients using chest X-ray images. Experiments were conduction on the Chest XR COVID-19 detection challenge dataset. Experimental results on a large-scale dataset showed that our late ensemble networks achieved 95.67%, 95.67%, and 95.77% accuracy, sensitivity, and specificity, respectively. The proposed approach was ranked 6th in Chest XR COVID-19 detection Challenge [1]

## Data Availability

Dataset is publicly available and listed in paper.

https://cxr-covid19.grand-challenge.org/

## Footnotes

1 https://github.com/RespectKnowledge/Chest-XR-COVID-19-detection*Deep - Learning*

2 https://www.worldometers.info/coronavirus/

